# Effect of preoperative educational counselling about routine elements of peri-operative care on patient’s experience through their first surgical journey: a randomised control trial

**DOI:** 10.1101/2023.01.09.23284099

**Authors:** Feba Susan Kurian, Santosh Balakrishnan, Joicy Els Jojo

## Abstract

**AIM:** To investigate whether pre-operative counselling about non-procedural elements of perioperative care improves patient’s overall surgical experience, through their first surgical journey

**OBJECTIVE:** To study the effect pre-operative counselling about non-procedural elements of perioperative care in a tertiary care centre, on the care experience of such patients.

**METHODS:** 62 Patients charted for their first-ever surgical procedure, were enrolled for the trial giving an Interventional group (A: n= 32) and control group (B: n = 30). Surgical experience was recorded 48 hours after surgery on regional-language questionnaire, developed & validated by an expert group. Data was compiled in Ms Excel and Statistical analysis was done using R software.

**RESULTS:** Mean scores were compared with paired T-test. Groups A and B had a mean experience score of 71.78 and 62.93 respectively (Mean difference (MD) 8.85; P <0.001). Mean scores in preoperative, intraoperative & post-operative domains were 33.31 vs 30.27 (MD 3.05; P <0.001), 14.47 versus 12.30 (MD 2.15; P <0.001) and 24.00 vs 20.37 (MD 3.63; P <0.001) for the groups respectively.

There was significant difference in surgical experience scores favouring the interventional group who recorded greater confidence and satisfaction with overall surgical experience and also separately for preoperative, intraoperative and postoperative periods.

**CONCLUSION:** Educating patients undergoing their first surgery about elements of perioperative care inherent to hospitalization for any surgery, has significant role in improving patients’ overall care experience in their surgical journey. Regularising such practice could improve overall surgical experience for patients.

## INTRODUCTION

Surgery can be daunting for patients, especially their first experience. Pre-operative counselling refers to an educational intervention before surgery with the aim of improving patient’s knowledge, health and outcome.^1^ This usually happens as part of informed consent. The patient is given information specific to the procedure and expected procedure related benefits, risks, safety & care arrangements available to ensure safe treatment.

Despite such procedure specific counselling being standard practice, patients often suffer peri-operative anxiety which overwhelmingly comes to define their experience with surgical care despite good treatment outcomes. Several international studies have looked into the effect of pre-operative counselling on individual aspects like anxiety^2,3^ length of stay^3^ and pain^2^. However, there is lack of data on the effect of pre-operative counselling about routine peri-operative hospital processes on the overall experience of the patient through their first every surgical care journey.

## AIM

To investigate whether pre-operative counselling about routine non procedural elements of perioperative care could improve patient’s overall surgical experience, through their first surgical journey.

## OBJECTIVE

To study the effect of a single episode of structured pre-operative counselling about routine non procedural elements of perioperative care in a tertiary care centre, on the personal experience of patients undergoing the first surgical treatment under anaesthesia in the main operating theatre.

## MATERIAL AND METHOD

After seeking clearance from the institutional ethics committee (IEC) and institutional review board (IRB), the trial was registered in Clinical Trial Registry -India (CTRI) and registration number was obtained. Patients charted for the first surgical procedure in their life, who satisfied our inclusion criteria were enrolled into the Randomised Control Trial.

Inclusion criteria: Inpatients admitted to surgical wards for general surgical & urology procedures at a tertiary care teaching hospital in the Ernakulam district of the south Indian state of Kerala with the following attributes

a. Age group:18-80 years
b. Charted for surgical procedures under general and spinal anaesthesia.

### Exclusion criteria

a. Patients undergoing major surgeries requiring post-operative intensive care other than immediate recovery from anaesthesia.
b. Patients undergoing minor surgeries under local anaesthesia.
c. Patients undergoing emergency surgical procedures.
d. Patients who have had previous surgical experience apart from obstetric procedures.

In the absence of a suitable questionnaire in the regional language, one was developed, validated by an expert committee of 9 members (including 6 doctors, 2 nurses, and a counsellor) and statistically validated by a qualified bio-statistician using Cronbach’s alpha. The questionnaire comprised 15 questions divided into three sections of 5 questions each to cover covering three periods of their surgical treatment journey; (domain 1: preoperative experience, domain 2: operation theatre experience, domain 3: postoperative experience) marked on a 5-point Likert scale. (Annexure 1)

Sample size (n=62) was calculated based on a pilot study on twenty patients accepting type I error α at 5% and type II error β at 20% and a standard deviation of 5. Patients were randomised into two groups by permuted block randomisation with allocation concealment using closed envelopes reducing recruitment bias.

Double blinding was achieved by ensuring delivery of experience assessment questionnaire 72 hours after surgery by an assessor blinded to the allocation. Equal time was spent speaking with subjects from both groups after group allocation to avoid subjective bias among the recruited subjects admitted to the same ward.

Statistical analysis of the data was done using R software

## OBSERVATIONS AND RESULTS

Male to female ratio was 59.4: 34.3 and 63.4: 36.5 in the test and control group respectively. (P<0.05) Majority of the participants in both groups had an educational qualification equivalent to a high school degree or higher degree. 56.2% of the participants in the test group and 63.3% of the participants in the control group underwent minimally invasive surgical and urological procedures (P<0.05).

The test and compared groups were compared for confounding factors such as influence of type of procedure & comorbidities and were found to be comparable in all other aspects other than the intervention under study.

The data of results was confirmed to follow normal distribution (Kolmogorov-Smirnov test). An independent sample T test was performed between the groups to check for significance of difference in the mean overall surgical experience score as well as mean of domain scores covering three periods of their surgical treatment journey facilitated by the design of the questionnaire. It was observed that surgical experience score of the test group was 71.78 (SD =3.2) and the control group was 62.93(SD = 6.44) with mean difference 8.85 [95% CI (6.21-11.49) P<0.001] (Table 1)

**Table 1:**
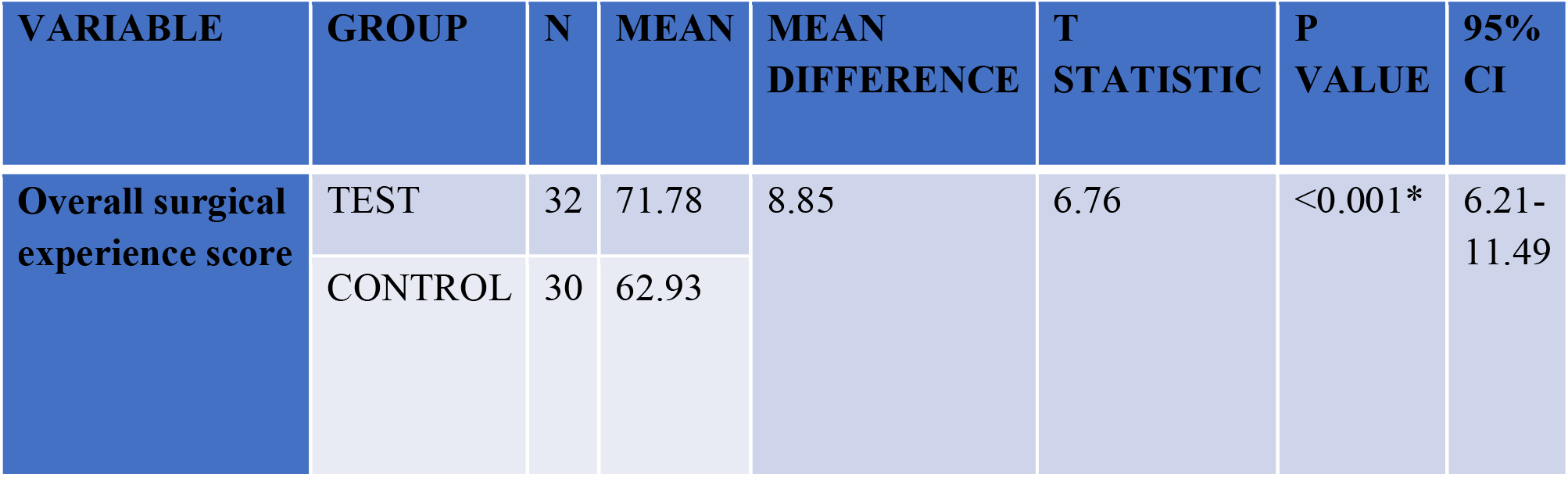
Mean surgical experience score-test versus control

| VARIABLE | GROUP | N | MEAN | MEAN DIFFERENCE | T STATISTIC | P VALUE | 95% CI |
| --- | --- | --- | --- | --- | --- | --- | --- |
| Overall surgical experience score | TEST | 32 | 71.78 | 8.85 | 6.76 | <0.001* | 6.21-11.49 |
|  | CONTROL | 30 | 62.93 |  |  |  |  |

We also observed that there was a significant difference in the average score between the control group and experimental group in all the 3 domain scores viz preoperative, Operation theatre and post operative experience score with mean differences of 3.05 [95% CI (1.43-4.67) P<0.001], 2.17 [95% CI (1.39-2.95) P<0.001] & 3.63 [95% CI (2.69-4.58) P<0.001] respectively. (Table 2)

**Table 2:** mean surgical experience score in 3 domains-test vs control

| VARIABLE | GROUP | N | MEAN | MEAN DIFFERENCE | T STATISTIC | P VALUE | 95% CI |
| --- | --- | --- | --- | --- | --- | --- | --- |
| Presurgical experience score | TEST | 32 | 33.312 | 3.05 | 3.76 | <0.001* | 1.43-4.67 |
|  | CONTROL | 30 | 30.27 |  |  |  |  |
| Experience score during surgery | TEST | 32 | 14.468 | 2.17 | 5.60 | <0.001* | 1.39-2.95 |
|  | CONTROL | 30 | 12.300 |  |  |  |  |
| Postsurgical experience score | TEST | 32 | 24.000 | 3.63 | 7.76 | <0.001* | 2.69-4.58 |
|  | CONTROL | 30 | 20.366 |  |  |  |  |

A subgroup analysis of the questionnaire response revealed that the sum of scores of the patients of agreement in principle to the positively worded questions (agree + strongly agree) and disagreement in principle to the negatively worded questions (disagree + strongly disagree) in control group agreed in principle to the statement that they felt confident in going ahead with the surgery with no disagreement in principle in either group to that question.

When similar scores were considered for strong agreement and strong disagreement to the same questions considered above, significant difference was observed between the 2 groups. 96.8% of the test group as against 66.7% of the control group strongly agreed to the statement that they felt confident in going ahead with the surgery.

## DISCUSSION

Surgery, however vital & well performed, takes an undeniably effect on the emotional health of a patient, which can in turn affect post operative physical recovery and compliance to future medical treatments.^4^

Patient satisfaction is as an important as the clinical outcome from surgery in terms of service delivery & planning. A key determinant of patient satisfaction is patient education.^5,6^ A common method of coping with an anticipated life event is by obtaining information which reduces the degree to which it is perceived as being stressful.^2^

The standard process of consenting for surgery focuses on treatment options, benefits versus risks of the proposed procedure, steps taken to ensure safe outcomes & expected recovery process including return to normal work & life.

While there has been recent emphasis on improving patient’s pre-operative, operative and post operative experience to improve satisfaction & aid recovery there was dearth of literature on the effect of preoperative educational counselling about routine elements of peri-operative care in improving improve a patient’s surgical experience. This study was therefore a novel attempt to investigate the effect of educating patients about routine elements of peri-operative care inherent to all surgical procedures, but often unaddressed, in improving patient’s overall surgical experience during their life’s first surgical treatment journey.

We observed from our study that there existed a significant difference in the surgical experience score between test and control groups (P<0.001). A significant difference in the average domain scores between the counselled group and uncounselled group was also noted in all the three questionnaire-defined treatment period domains (preoperative, Operation theatre and postoperative experience domains). A statistically significant difference was thus observed in the surgical experience score between the groups in favour of the test group validating the effectiveness of the study intervention.

The sub-group analysis of the questionnaire response observed that the sum of overall agreement in principle (agree + strongly agree) and disagreement in principle (disagree + strongly disagree) between both groups were not significantly different. This in all possibility reflects the effect of the robust surgical consenting process which ensured that all the patients were certain beyond doubt that they need to undergo surgery with all its inherent benefits and risks even though they had less than maximal confidence in their experience reflected in the postop questionnaire score.

However the statistically significant difference in favour of the test groups when responses of strong agreement or strong disagreement was considered clearly reflects the influence of the counselling intervention in boosting confidence of the patients to certainty as also reflected by other studies that offer similar evidence for the positive effects of preoperative counselling on different variables post operatively.^7,8,9,10,11,12,13^

Multiple studies have proven that satisfied patients have better health outcomes because they tend to the obey doctor’s advice, refrain from malpractice litigations, comply with treatment regimens, attend follow up appointments, and ask for medical advice when required.^14,15,16^ A meta-analysis of 68 studies undertaken by Hathway et al indicates that patients who receive preoperative education have 20 percent favourable postoperative outcomes of physiological variables, (length of stay, sedatives used, recovery, complications) and psychological variables (observed ratings of cooperation, scores of self-reported anxiety inventories, etc) compared to those patients who did not receive preoperative education.^8^ These results are comparable to our study wherein in we observed favourable outcomes among the test population in terms of various preoperative, intraoperative and postoperative elements.

To balance our conviction based on our observations we would like to also highlight a few studies that show negative or no effect of pre-operative counselling on patient outcomes.^17,18^ They report difficulties experienced in counselling or patient’s inability to understand or use the information provided as reasons for the contrary results. The high level of literacy in the state of Kerala, delivery of structured information in the patient’s mother tongue as well as use of a purpose designed & validated questionnaire in the regional language allowed us to to minimize the effect of these confounding factors experienced by the aforementioned researchers, in the present study.

## CONCLUSION

Preoperative educational counselling about routine elements of peri-operative care in addition to routine surgical consent process significantly improved the patient’s experience through their first surgical journey.

We believe routine introduction of such counselling for patients regarding peri-operative care could serve to improve the patients overall surgical experience ánd in turn translate to higher level of confidence among patients to seek treatment when needed without fear, thus improving outcomes.

## Data Availability

ALL DATA PRODUCED IN THE PRESENT STUDY ARE AVAILABLE UPON REASONABLE REQUEST TO THE AUTHORS

## SUPPORTING DOCUMENTS

### 1. CASE STUDY FORM

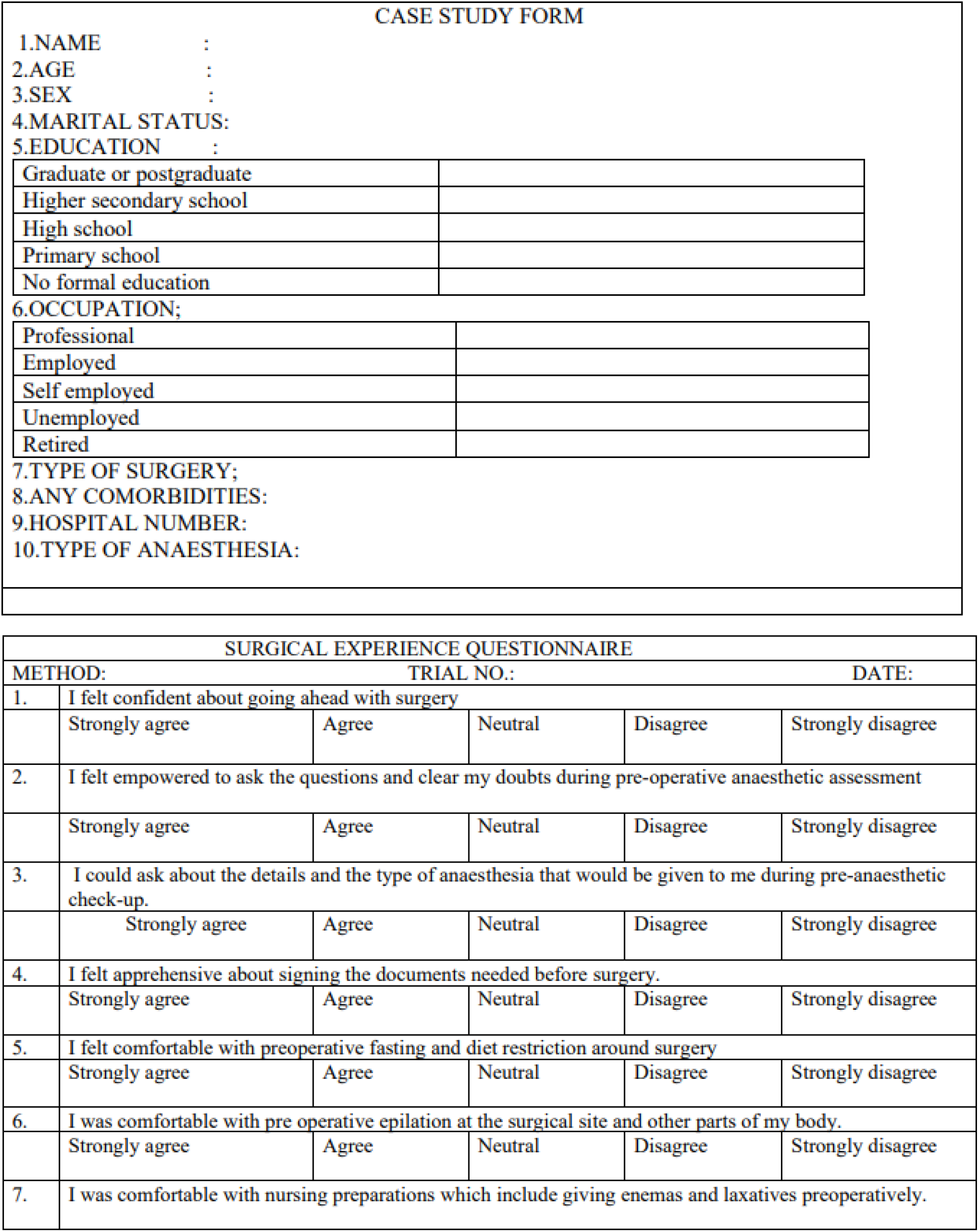
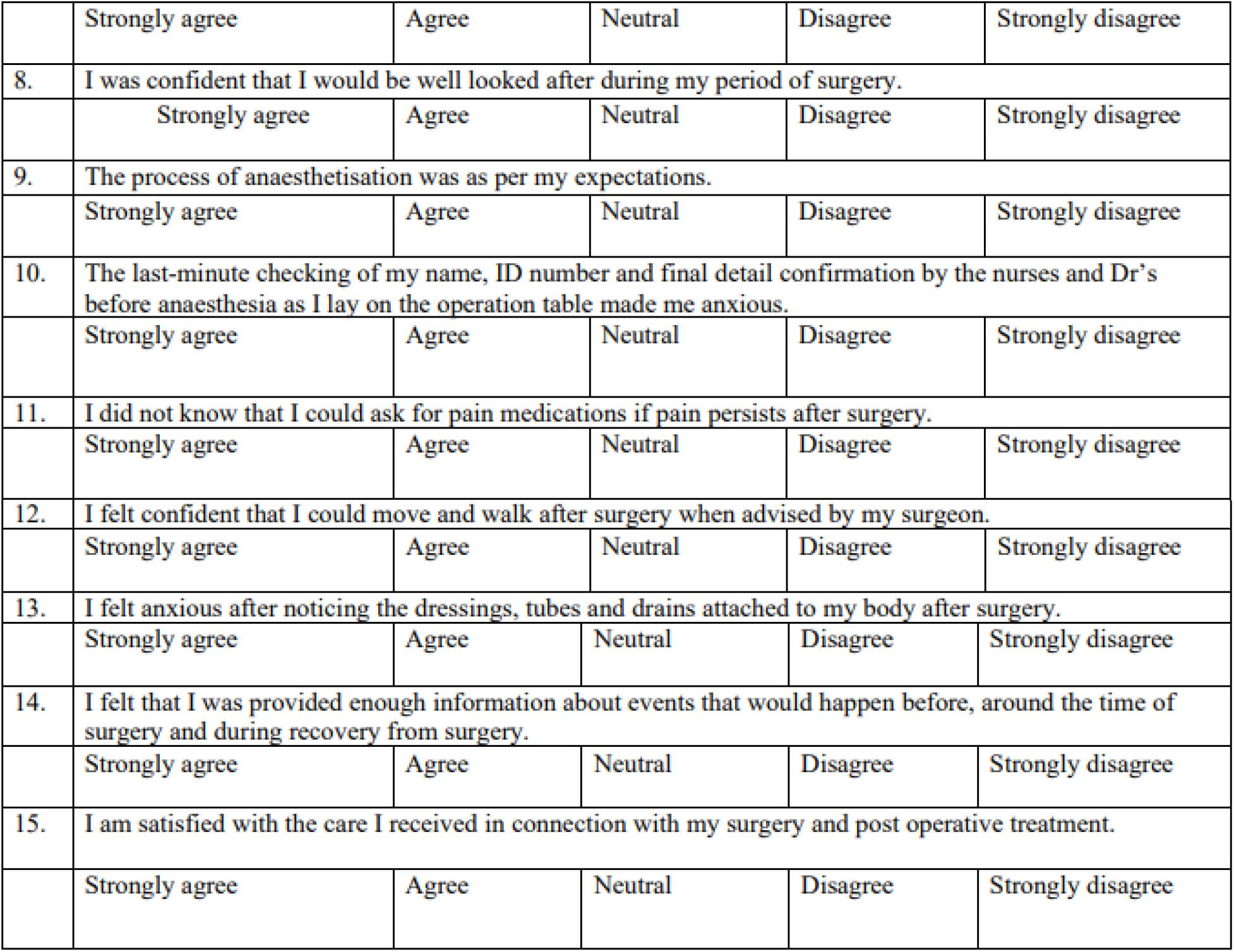

1.CASE STUDY FORM
| CASE STUDY FORM |
| --- |
| 1.NAME : |
| 2.AGE : |
| 3.SEX : |
| 4.MARITAL STATUS: |
| 5.EDUCATION : |
| Graduate or postgraduate |
| Higher secondary school |
| High school |
| Primary school |
| No formal education |
| 6.OCCUPATION; |
| Professional |
| Employed |
| Self employed |
| Unemployed |
| Retired |
| 7.TYPE OF SURGERY; |
| 8.ANY COMORBIDITIES: |
| 9.HOSPITAL NUMBER: |
| 10.TYPE OF ANAESTHESIA: |

| SURGICAL EXPERIENCE QUESTIONNAIRE |  |  |  |  |  |
| --- | --- | --- | --- | --- | --- |
| METHOD: |  | TRIAL NO.: |  | DATE: |  |
| 1. | I felt confident about going ahead with surgery |  |  |  |  |
|  | Strongly agree | Agree | Neutral | Disagree | Strongly disagree |
| 2. | I felt empowered to ask the questions and clear my doubts during pre-operative anaesthetic assessment |  |  |  |  |
|  | Strongly agree | Agree | Neutral | Disagree | Strongly disagree |
| 3. | I could ask about the details and the type of anaesthesia that would be given to me during pre-anaesthetic check-up. |  |  |  |  |
|  | Strongly agree | Agree | Neutral | Disagree | Strongly disagree |
| 4. | I felt apprehensive about signing the documents needed before surgery. |  |  |  |  |
|  | Strongly agree | Agree | Neutral | Disagree | Strongly disagree |
| 5. | I felt comfortable with preoperative fasting and diet restriction around surgery |  |  |  |  |
|  | Strongly agree | Agree | Neutral | Disagree | Strongly disagree |
| 6. | I was comfortable with pre operative epilation at the surgical site and other parts of my body. |  |  |  |  |
|  | Strongly agree | Agree | Neutral | Disagree | Strongly disagree |
| 7. | I was comfortable with nursing preparations which include giving enemas and laxatives preoperatively. |  |  |  |  |
|  | Strongly agree | Agree | Neutral | Disagree | Strongly disagree |
| 8. | I was confident that I would be well looked after during my period of surgery. |  |  |  |  |
|  | Strongly agree | Agree | Neutral | Disagree | Strongly disagree |
| 9. | The process of anaesthetisation was as per my expectations. |  |  |  |  |
|  | Strongly agree | Agree | Neutral | Disagree | Strongly disagree |
| 10. | The last-minute checking of my name, ID number and final detail confirmation by the nurses and Dr's before anaesthesia as I lay on the operation table made me anxious. |  |  |  |  |
|  | Strongly agree | Agree | Neutral | Disagree | Strongly disagree |
| 11. | I did not know that I could ask for pain medications if pain persists after surgery. |  |  |  |  |
|  | Strongly agree | Agree | Neutral | Disagree | Strongly disagree |
| 12. | I felt confident that I could move and walk after surgery when advised by my surgeon. |  |  |  |  |
|  | Strongly agree | Agree | Neutral | Disagree | Strongly disagree |
| 13. | I felt anxious after noticing the dressings, tubes and drains attached to my body after surgery. |  |  |  |  |
|  | Strongly agree | Agree | Neutral | Disagree | Strongly disagree |
| 14. | I felt that I was provided enough information about events that would happen before, around the time of surgery and during recovery from surgery. |  |  |  |  |
|  | Strongly agree | Agree | Neutral | Disagree | Strongly disagree |
| 15. | I am satisfied with the care I received in connection with my surgery and post operative treatment. |  |  |  |  |
|  | Strongly agree | Agree | Neutral | Disagree | Strongly disagree |

### 2. COUNSELLING PROFORMA

1. Preoperative investigations. Explanation about preoperative evaluation which includes measuring of height, weight and vital signs, reviewing medication list with recommendations for how to take medications prior to surgery, reviewing medical, surgical and family history, physical examination, additional preoperative testing following your exam if deemed necessary. This may include, blood or urine tests, Xrays or an electrocardiogram.
2. Explanation about preanesthetic check process. Enlightening the patient about arrival of anaesthetist and the process which includes taking affirmative history, estimation of blood haemoglobin, blood loss estimation, blood availabity. Educating about different types of anaesthesia [spinal /epidural/general], analgesics available during post operative Period and their right to ask for additional doses while in pain. We would also empower them to clarify their doubts and inform any known drug allergies or use of drugs during preoperative anaesthetic assessment. We would also inform them about the process of giving consent prior to induction of anaesthesia would sensitize them about the arrival of anaesthetist who would decide on the type of anaesthesia after taking the interest of patient into consideration, after explaining the pros and cons of the procedure. Information about consent if blood transfusion is required is given to the patient.
3. Preoperative physical preparations. Information about preoperative epilation and its role in reducing surgical site infections will be explained to the patient. The patient would also be made aware of the chance of epilation in other areas away from the surgical site if necessary. Bowel preparations with laxatives or enemas will be explained to the patient.
4. Perioperative dietary restriction. The patient is made aware of the reasons for having an empty stomach before any surgery or procedure that require anaesthesia such as to prevent nausea and to keep any food or liquid from entering into lungs.
5. Explanation about processes in the morning of surgery. A brief idea about the processes in the morning of surgery will be explained to our patient right from arrival of trolly, preoperative epilation, verification of the details at gate, WHO callout and placing the patient on the OT table before anaesthesia, insertion of IV canula, premedications, induction of anaesthesia is given to the patient.
6. Process of going through anaesthesia. Information about types of anaesthesia in general, sedatives and analgesics, insertion of IV canula, and basic information regarding induction of anaesthesia is given to the patient.
7. Explanation about immediate postoperative recovery and transfer. A brief idea about recovery and postoperative ICU setup, Oxygen supply through tubes in nose or mask is given to the patient. Continuous monitoring of BP and vitals will be done. Empowerment to ask for analgesic while in pain is given more times to the patient
8. Information about tubes, drains, and dressings. The patient will be counselled about the possibility of finding tubes or drains coming out of the body to assist with prevention of collecting fluid, blood or pus at the surgical site. They will also be counselled about the potential need for urinary catheter and its rationale. They are also informed that initial dressing may be large and it has no bearing on the intraoperative success or outcome of the operation.
9. Confidence to move and walk with the advice of surgeon The patient is made aware that earlier he/she get out of bed and start walking, eating and drinking after the operation the more better. Rehabilitation programs and other methods of recovery are introduced. Empowering patients to move and walk as advised by the surgeon and reassure them that no harm will come from such activity as advised by the treating doctor.

